# Background and Foreground: Connections & Distinctions when Health Professions Faculty Teach Both Interprofessional Collaborative Practice and Quality Improvement – A Case Study

**DOI:** 10.1101/2024.06.11.24308788

**Authors:** Katherine Stevenson, Johan Thor, Marcel D’Eon, Linda A. Headrick, Boel Andersson Gäre

**Affiliations:** the Jönköping Academy for Improvement of Health and Welfare, at the School of Health and Welfare, Jönköping University, Jönköping, Sweden; College of Medicine, University of Saskatchewan, Saskatoon, Saskatchewan, Canada; School of Medicine, University of Missouri, Columbia, Missouri, United States

## Abstract

Despite decades of effort, programs continue to struggle to integrate competencies related quality improvement (QI) and interprofessional collaborative practice (ICP) into health professions education. Additionally, while QI and ICP may seem intuitively linked and there exists some examples of a coordinated approach, the literature regarding competencies, including knowledge, skills, and attitudes (KSAs), is still largely focused on QI and ICP as separate fields of knowledge and practice. This study explored distinctions and connections between quality improvement (QI) and interprofessional collaborative practice (ICP) competency domains in health professions education. The authors used a qualitative case study approach with an instrumental case, i.e., the University of Missouri-Columbia (MU), where QI and ICP were intentionally integrated as part of core curricula in health professional schools and programs. Eleven faculty members from medicine, nursing, pharmacy, and health care administration participated in interviews exploring their teaching choices in either classroom or clinical settings. Study participants defined the goal of teaching QI and ICP as enabling learners to deliver safe and patient-centered care and described the knowledge and skills required for QI and the attitudes and skills required for ICP. Furthermore, they described the relationship between QI and ICP as one mediated by systems thinking, where ICP is backgrounded as a critical pre-requisite and QI is foregrounded as a vector for developing interprofessional competencies. The MU case elucidates the potential synergies that occur when faculty address quality improvement and interprofessional collaborative practice competencies with an integrated approach that leverages connections, while also respecting distinctions. For health professions education programs looking to improve the effectiveness and efficiency of their curricular approach to these fields, it may be fruitful to consider ICP as background and QI as foreground, remembering that without each other, ICP risks losing meaning and QI risks losing impact.

## Background

As concerns over health care quality, complexity, fragmentation, and cost have grown increasingly pertinent, two content areas have emerged as integral in health professions education: interprofessional collaborative practice (ICP) and quality improvement (QI).^1^ Despite decades of effort to integrate these content areas into health professions education, many programs continue to struggle. And while there are some examples of a coordinated approach, the literature regarding competencies, including knowledge, skills, and attitudes (KSAs), is still largely focused on ICP and QI as separate fields of knowledge and practice.

Much of the groundwork for our understanding of both interprofessionalism and collaboration can be traced back to a seminal work by social worker Dr. Rosalie Kane, *Interprofessional Teamwork*.^2^ Interestingly, a focus on ICP was initiated prior to Dr. Kane’s publication, as early as the 1960s.^3^ Globally, ICP and related educational initiatives were marked by the development, in 2002, of what is now the most widely quoted definition of interprofessional education (IPE): “occasions when two or more professions learn with, from and about each other to improve collaboration and the quality of care.”^4^ Another key landmark was the release of the Framework for Action on Interprofessional Education & Collaborative Practice by the World Health Organization where they reaffirmed the CAIPE definition of interprofessional education and defined ICP as “multiple health workers from different professional backgrounds provide comprehensive services by working with patients, their families, carers and communities to deliver the highest quality of care across settings.”^5(p13)^ Despite decades of focused research and effort, one of the hallmarks of ICP and IPE is that there is still wide variation in terminology and definitions, with a call for standardization coming as recently as 2022. ^6,7^ For the purpose of this work, which focuses on both the academic classroom environment and the care delivery environment, we use ICP education to refer to any educational intervention looking at improving collaborative practice (as defined by the WHO), including IPE (as defined by CAIPE). In the early 2000s, the Institute of Medicine’s landmark reports,^b^ *To Err is Human* and *Crossing the Quality Chasm*, heightened the focus on QI in healthcare delivery.^8,9^ We have used the definition of QI from Batalden and Davidoff, i.e. QI is the “combined and unceasing efforts of everyone—healthcare professionals, patients and their families, researchers, payers, planners and educators—to make the changes that will lead to better patient outcomes (health), better system performance (care) and better professional development (learning)”.^10(p2)^ We can trace initial efforts to introduce QI as a topic for health professions education as far back as the 1990’s, following the move to “managed care” in the United States and a shift in focus from quality assurance to quality improvement. ^11–13^ Early efforts included the Institute for Healthcare Improvement (IHI) Interdisciplinary Professional Education Collaborative, which deliberately and clearly linked QI and ICP with a focus on teams as the process and improvement as the content of the learning.^14^ In the late 90s, the IHI also defined Eight Knowledge Domains for Health Professional Students, which have greatly influenced educators working in this space.^15^

Despite early signals of connection across these two fields, the teaching, research, and practice communities appear to have pursued separate approaches to QI education and ICP education, creating two distinct bodies of knowledge. The QI education field has mostly focused on topic– and profession-specific curriculum frameworks and competency statements, without a broadly embraced, unifying and interprofessional statement or framework.^16,17^ Work in ICP has, perhaps naturally, been less profession-specific and includes national level descriptions of competencies to help guide curriculum development, but does not fully integrate QI.^18,19^

For example, the Canadian interprofessional competencies are inclusive of QI, but treat it as a background consideration alongside contextual factors and complexity.^19^ The American *Core Competencies for Interprofessional Collaborative Practice* go further to include QI as one explicit competency in the “teams and teamwork” general competency domain, but stop short of providing specifics.^18^ One notable exception is the Institute of Medicine’s report *Health Professions Education: A Bridge to Quality*.^20^ The report recommended a set of five core competencies common to all professions, including the ability to provide patient-centered care, work in interdisciplinary teams, and utilize informatics. It goes so far as to highlight the interaction amongst the competencies, which are inclusive of both QI and ICP related KSAs. Still the recommended competencies are not fully operationalized across all aspects of clinical practice and health professions education, with the nursing profession providing the only example we can find in the form of Quality and Safety Education for Nurses.^21^

In reviewing education-related literature in both fields, it appears as though ICP proponents often see quality as a byproduct of effective teamwork, while QI advocates primarily see teams as prerequisite to achieving optimal quality.^22–32^ While the two fields acknowledge each other, each appears to take for granted the specific evidence-base of the other. For example, scholars have been developing the science around quality improvement drawing on particular theories and tools, but these are often overlooked in ICP education literature.^33–35^ Even effective collaborative clinical teams may not achieve their potential, in terms of quality of care and outcomes, without using or applying the QI evidence base. On the other hand, there are social and behavioral sciences underpinning the field of ICP that are often oversimplified or even ignored in QI education literature.^17,36–38^ For example, one cannot assure effective collaboration simply by making groups working on quality improvement projects multiprofessional;^c^ instead attention must be paid to the principles of group dynamics and issues of professional culture and power.^40–45^

While there are advantages to maintaining a separation between the fields of QI education and ICP education, including academic recognition tied to visibility and consistency of identity, such separation can generate confusion and difficulties for academic and clinical educators working on curricular change.^46^ More importantly, such a division misses the potential synergies between ICP and QI and is a barrier to a more integrated, and thus possibly more effective and efficient, approach to improving education and care.

To better understand what QI and ICP competency development looks like when integrated, an example can help. As a university community working explicitly on both QI and ICP education for both the pre– and post-licensure learners, particularly in the 10 years between 2003 and 2013, the University of Missouri-Columbia (MU) offers a possibility for rich learning. Of particular interest to us are the perspectives of faculty members. Faculty members are crucial actors as they take program theories such as key frameworks and competency statements, and translate them into practice in classrooms or clinical education situations.^47^ Exploring faculty members’ beliefs and experiences with ICP education and QI education in practice provides an opportunity to refine our understanding of the relationship, unique features, and possible linkages between the competency domains related to each field.

The aim of this paper is to highlight potential synergies by exploring and describing connections and distinctions between QI and ICP competency domains from the perspective of faculty members teaching an interprofessional quality improvement curriculum.

## Methods

We used an instrumental case study^d^ approach underpinned by an interactive research model, with the case being the faculty’s experience with integration of QI and ICP at MU from 2003-2013. In interactive research we pursue a threefold task: contribute meaningfully to practical concerns, create new knowledge, and learn together as research and practice systems interact through the course of a project.^48^ As with other forms of collaborative research, the distance between the researched and the researchers is often diminished; specifically, LH, a faculty member at MU, did not participate in data collection or analysis, but did contribute to the interpretation and writing process.

## Context

The University of Missouri-Columbia (MU) is one of four campuses making up the University of Missouri System. With over 30,000 enrolled students, MU, working collaboratively with the other three University of Missouri campuses, provides a comprehensive suite of health programs, including medicine, nursing, pharmacy, rehabilitation sciences, and health care administration. In 2003, MU introduced a combined interprofessional-quality improvement education initiative involving both academic and clinical faculty.^49^ Over the ten-year period in focus, they progressively developed curricular improvements to teach health professions learners at all stages of formation in an integrated way both how to work together in interprofessional teams and how to continuously improve their work through application of quality improvement methods and tools. Not only have they developed and evaluated their innovations in teaching, but they have also published extensively. ^49–56^ The MU School of Medicine has been recognized for its efforts with the Learning Health System Challenge Award.^57^ As elsewhere, research in this area at MU has generally focused on the learner experience with little attention paid to faculty perspectives.

## Participants

We used judgment sampling, guided by MU academic program leaders, to invite academic and clinical faculty participants from a variety of backgrounds who had experience leading and/or teaching in the interprofessional quality improvement education program from 2003-2013. ^58^ This yielded 11 faculty members, representing Medicine (6), Nursing (3), Pharmacy (1), and Health Care Administration (1). Four of these participants had roles primarily on the academic side and seven on the clinical side. Seven of them (four primarily academic and three primarily clinical) were also connected to the Center for Health Quality at the University of Missouri, which bridged the academic and clinical learning environments. Interview participants (n=11) were active in QI education and ICP at either, or both, the undergraduate (n=7) and postgraduate level (n=7).

## Procedure for data collection and analysis

Each faculty member participated in an initial one-on-one interview with KS (the first author) using a semi-structured interview guide, field-tested elsewhere (not published), from a case-study perspective.^59^ The focus of the interview was on the what, the how, and the why of teaching choices related to QI and ICP. Where appropriate and feasible, KS conducted a second interview (n=6) using a modified stimulated-recall technique.^e^ Stimulated-recall is a collection of strategies designed to uncover thought processes and beliefs by anchoring an interview in an observation of practice.^60^ In this study, we used stimulated recall as a reflective practice tool with respondents to “make explicit and articulate the thinking, knowledge, theories and beliefs that guided their teaching practice” and inquire into deeper reflection on action.^60(p.290)^ The object most commonly used to stimulate recall is video recording of an event, e.g., a teaching session. However, due to concerns that recording teaching in this context might interfere with student learning and/or not be feasible due to teaching modes (e.g., simulation), we modified the technique by using notes taken by KS during observation of teaching sessions and learning objects, i.e. teaching materials and student handouts, as the stimulus. We collected all data in January 2013 and used QSR NVivo 10™ software to support analysis of transcribed data from the initial and stimulated-recall interviews.

We approached data analysis and synthesis from an abductive perspective, where data are analyzed through a lens of “extensive familiarity with existing theories.” ^61(p173)^

Abductive analysis offers the potential to propose possible solutions to challenging problems using a “a back-and-forth process between the research evidence and considerations of theory.”^62,63(p305)^ In abduction, we still engage in familiar processes for qualitative data collection and coding, but are “sensitized to what is novel vis-a-vis theory.”^64(p2)^ KS coded the data using thematic content analysis to form categories and subcategories.^65^ The categories and subcategories were iteratively refined with relevant theories in mind, including Bloom’s taxonomy and descriptions of both QI and ICP competencies.^15,16,18,19,66,67^ KS then shared the results with six respondents in a member check conference call and provided an opportunity for them to reflect on to what extent the findings resonated with them. Two additional respondents provided written feedback. Feedback from respondents resulted in overall confirmation of the categories and moderate refinement of the subcategories. Finally, KS discussed the analysis with all co-authors to ensure consistency of the process as well as the coherence and validity of the findings.

## Findings

Respondents described 1) the goal of quality improvement and interprofessional collaborative practice, 2) knowledge, skills, and attitudes required for improving quality (knowledge and skills) and effective interprofessional team collaboration (attitudes and skills), and 3) the relationship between quality improvement and interprofessional collaborative practice. Table 1 overviews a summary of categories and sub-categories. ***The goal of quality improvement and interprofessional collaborative practice*** Faculty members described a core feature of the education program as keeping the learners focused on the various Institute of Medicine (IOM) aims for quality and on addressing gaps in the current system, i.e., safety, timeliness, efficiency, effectiveness, equity, and patient-centeredness. Paying attention to all six IOM aims, respondents emphasized the need for students and other trainees to learn to provide patient-centered care. While some teaching materials implied patient-centeredness was at the core of the work, faculty members reported that they placed an often-equal emphasis on safety.

**Table 1.** Summary of Categories and Sub-Categories.

|  | Category | Sub-Category |
| --- | --- | --- |
| The Goal | IOM aims for quality & safety | Priority to aims of Patient-Centered & Safe |
|  |  | Focus on the Interaction between the Aims |
| Core Knowledge (Kn) & Skills (Sk) for Quality Improvement | Analysing problems, processes, and systems | Exploring contributing factors (Sk) |
|  |  | Process or Flow Mapping (Sk) |
|  |  | Understanding Variation (Kn) |
|  |  | Measurement (Sk) |
|  | Applying effective change management strategies | Starting Small & Scaling Up (Sk) |
|  |  | Engaging Others (Sk) |
|  |  | Calculating Effort vs Yield (Sk) |
|  |  | Focusing on Implementation (Sk) |
|  |  | Bridging Clinical and Business Aims (Sk) |
|  |  | Specific QI Approaches (Kn) |
|  | Leveraging information technology | Managing Data (Sk) |
|  |  | Presenting Ideas (Sk) |
| Core Attitudes (Att) & Skills (Sk) for Effective Interprofessional Collaboration | Individual Orientation | Curious (Att) |
|  |  | Mindful (Att) |
|  |  | Self-Aware (Att) |
|  |  | Humble (Att) |
|  | Team Orientation | Inclusive (Att) |
|  |  | Interdependent (Att) |
|  |  | Reflective (Sk) |
|  |  | Psychologically Safe (Att) |
|  |  | Problem-Solving Focus (Att) |
|  | Constructive Interaction | Moderating Psychological Size (Sk) |
|  |  | Speaking Up (Sk) |
|  |  | Receptivity (Sk) |
|  |  | Sharing Knowledge (Sk) |
|  |  | Sharing Decision Making (Sk) |
|  |  | Resolving Conflicts (Sk) |
|  |  | Negotiating Culture (Sk) |
| The Relationship between QI and ICP | ICP as Background & Pre-Requisite | Providing Context |
|  |  | Solving Complex Problems |
|  |  | Creating Sustainable Solutions |
|  |  | Meeting the Needs of the Whole Person |
|  | QI as Foreground & Vector | Vector for ICP |
|  |  | Provides a Purpose for ICP |
|  |  | Limits of Individual QI |
|  | System's Thinking as a Mediating Interface | Core Skill in QI |
|  |  | Context for ICP |

> *I mean to me they’re both at the center and your first priority is the patient and keeping the patient safe and those sort of go hand in hand. So, on the diagram even though the patient is at the very center that just symbolizes the fact that in everything we do the patient has to be the center…they’re almost intertwined. (K)*

Many of the faculty members recognized the importance of seeing the interactions among the six IOM aims, in that they aren’t isolated, but often work synergistically.

> *So, what we’re trying to do is, on those patients at risk for discharge issues, move the process up [i.e., start discharge planning on admission]. So that’s clinical, but it’s the right thing to do because it’s good care [effective]. It’s less expensive. It’s less wasteful [efficient]. Patients are happier [patient-centered]. (E)*

This synergy was especially evident to them when teaching clinical skills in the care setting.

## Core knowledge and skills for improving quality

In addition to developing clinical skills, respondents emphasized the need for learners to develop improvement capability, specifically leveraging information technology to support their efforts in the following areas: analyzing problems, processes, and systems as well as applying effective change management strategies.

When teaching about process and system analysis, faculty focused on specific tools, e.g., process mapping and contributing factors analysis using fishbone diagrams,

> *…we introduce them to the concept of being able to look at a case like an adverse event or in a non-judgmental way and as a team to map out of the flow of care and look at what steps occurred in the care delivery (K)*

> *We have them do…a fishbone diagram… I think [fishbones and process maps] are some of the easiest …and most helpful tools in quality improvement and are ones that are often not done. And I use them all the time. (F)*

They also emphasized measurement and understanding of variation, specifically, how to choose measures and gather, display, analyze, and interpret data.

> *…you have to have data, you have to have some basic understanding of how to interpret data in a meaningful way, understanding of…variation (D)*

> *I have a lecture called from anecdote to measurement and we simply don’t allow anecdotes… [I teach] them that data [is] their voice. (C)*

Applying effective change management strategies was not about using any one particular theory or method of change, but rather an amalgamation of practical and pragmatic tactics. These included starting small and then scaling up, engaging others, understanding how to analyze change ideas from the perspective of effort versus yield, and focusing on implementation strategies rather than simply generating ideas.

> *…we’ve proven that if we do this in six rooms, here’s the increase in patient satisfaction…, well let’s try it in twenty patient rooms… let’s try it on a whole wing, a whole floor. (G)*

> *Really what you’re doing in improvement is you’re selling somebody on the idea of changing their behavior. (E)*

> *Do you have the resource to fix it? And what you’re spending to fix, is it really worth fixing at that point? So, you’re trying to get low cost of resources and high yield of the change you can implement. (G)*

> *You realize of course that… 80 percent of improvement work is actually implementing the brilliant design you just came up with, and what makes a difference between the superstar and the people who are just smart are the people who can do the implementation piece. (A)*

More senior and post-licensure learners were also challenged to develop skills in bridging the clinical and business aims of the organization,

> *Quality is not the domain of the clinician, and I also approach it from the standpoint that clinical stuff is not the exclusive domain of the clinician because the administrators, the support systems that support it, actually are part of the clinical quality. (E)*

While the respondents described some degree of explicit teaching around Plan-Do-Study-Act (PDSA) cycles, they also voiced a reticence to focus too much on specific improvement approaches, i.e., Model for Improvement, Lean or Six Sigma,

> *They learn about PDSA Cycles … I’m not sure they’re ready to learn about Lean or Six Sigma. That doesn’t relate in their eyes as directly to the patient as some of these other things do. (J)*

Finally, several times faculty noted the importance of learners having or developing skills with information technology. Use of spreadsheet software, i.e., Excel, was seen as critical for data management and display,

> *And they had some examples, so now here you have this group of numbers; how do you order the numbers from top to bottom? How do you do an average? Now you have this set of things… make a little graph out of it. (F)*

and effective use of presentation software was linked to strategies in change management, i.e. the ability to “sell” your ideas and garner support,

> *And the final presentation is in the auditorium, it’s business dress, you come in, you have your [slides], …we invite… administration, faculty members. (G)*

### Core skills and attitudes for effective interprofessional team collaboration

In terms of teaching interprofessional team collaboration, respondents discussed Individual Orientation, Team Orientation, and Constructive Interaction. Individual Orientation is about the attitudes health professional students need to thrive in an interprofessional and collaborative environment: curious, mindful, self-aware, and humble.

> *Their favorite thing to do is to explore roles and responsibilities and learn about each other and you know they have no idea what the other person does… (K)*

> *We’re trying …to try to show them that as humans we can’t be perfect. We can’t ever give perfect care, … (I)*

> *The first competency is an understanding that …for you to give high value work, high quality work, you need to constantly be looking at your own practice and how you do things… (F)*

> *…they always need to question the boundaries of their [own] knowledge. (J)*

Additionally, faculty described the orientation of effective teams as being inclusive, interdependent, reflective, psychologically safe, and having a focus on problem solving.

> *They have a team huddle and everybody including the housekeeper goes to the huddle. And so, it’s first-name basis, everybody… (I)*

> *It is extraordinarily foolish to train just a single group of professionals to look at a problem in healthcare… tell me the problem that doesn’t involve at least two different disciplines and three other administrative units. (E)*

> *… they debrief, what went well and what could be improved. … And so, the team actually learns to critique themselves… it’s a little uncomfortable initially. People can say what we did well but what we could improve, it’s a lot tougher… (C)*

> *… the basic underpinning for the team is psychological safety. So that would be getting to know your teammates and feeling comfortable…to speak up. (I)*

Constructive interaction was described from four perspectives. First, that individuals within teams are able to moderate their psychological size, i.e., being attentive to their own authority and create space for others to engage,

> *If you watch people interact, you’re able to see the psychological size. So, a president at the university and an assistant professor have a different psychological size when they enter into a conversation. I’m looking for more parity among all the members of the healthcare team. (J)*

When that space is created, another aspect of constructive interaction is for individuals to have the courage to speak up and use their voice,

> *Can you talk to the other group, and can we break down the walls of being afraid because you think they’re smarter or you think they’re whatever. (B)*

Moderating psychological size and speaking up, allows for teams to be able to effectively share knowledge,

> *…it’s not about the knowledge that you have, it’s how you convey that knowledge to the other people that you’re working with. (H)*

and engage in shared decision-making,

> *…how to come to some shared decision making and incorporate somewhat divergent opinions. (K)*

When describing the concept of “team” in their teaching, faculty included patients and family members as full members, which emphasized again their focus on patient-centered care. Participants noted two skills that were important but not receiving enough emphasis in the curriculum as a whole: conflict resolution,

> *one of the gaps is …conflict resolution from a standpoint of…so you have a patient, and the nurse is really concerned about them, and you don’t see that there’s anything wrong …we need something further…where we give them a little bit more in the way of strategies to deal with things like that. (K)*

and negotiating professional and organizational culture,

> *I do think that we teach students more about the cultural competency of [working with] patients instead of …how much culture and context plays a role [professionally]…we expect of our students to understand the culture without even explaining it to them. (H)*

### The relationship between quality improvement and interprofessional collaborative practice

We specifically asked faculty to reflect on the relationship between QI and ICP. Generally, faculty described ICP as background and QI as foreground. ICP was described as a context for work, learning, and improvement,

> *…interprofessional education is more the context, so we sort of set that up as the context for their quality improvement and learning and work. (D)*

Additionally, ICP was seen as pre-requisite to solving complex clinical and system problems,

> *…the relationship to me is that not only is it common ground but that it would be hard to even try and solve a problem if you were just doing it on your own… (E)*

creating sustainable solutions,

> *[The] want to solve this problem and they work together on solving it. They demonstrate it’s solved. It’s probably going to be a sustainable solution that they’ve pieced together…(C)*

and meeting the needs of the whole person,

> *I think that to do good quality improvement you have to think about the experience of the patient. You have to see their illness through their eyes, or to see their experience through their eyes. Which means that it’s not just about medicine…it’s about culture…it’s about religion…it’s a prerequisite to meet the needs of the whole patient. (J)*

QI, on the other hand, was seen as a vector for ICP. Where learners might struggle to understand or have the insight that they didn’t already know how to be effective collaborators, QI was the vehicle to bring them together, inviting them explicitly with a problem to solve and implicitly with developing and honing the skills to do it together,

> *So, a vector for IPE would basically be the content area, so …quality and safety training might be a vector, end of life care might be a vector. In this case we used error disclosure training as a vector and… there is an explicit curriculum, so we’re going to learn about end-of-life care, but there’s an implicit curriculum as well which is we’re going to learn about teamwork and collaboration. (A)*

QI was also seen as providing a purpose for collaboration,

> *Because if you put an interprofessional team together and they don’t understand quality improvement, they can actually just be a very dysfunctional interprofessional team and that’s not necessarily going to produce good care. (A)*

While QI was acknowledged as being a unique person’s task on occasion, faculty emphasized the limitation of an individualistic approach, pointing to the complexity of most problems facing healthcare,

> *And so, you can’t really solve quality improvement problems without people from different professions… it’s not even something that I see that you can do, professionally. It’s hard to solve healthcare system problems in a silo… And so, I guess, yeah, the relationship to me is that not only is it common ground but that it would be hard to even try and solve a problem if you were just doing it on your own. (H)*

Additionally, a skill described by faculty as “systems thinking” emerged as a mediating or bridging category. Understanding, analyzing, and being able to create change in systems was a focus within the teaching of improvement skills, and inherent in many of the tools of improvement, e.g., fishbone diagrams and process mapping,

> *…the systems thinking is there, that people naturally start looking at things …as processes rather than as individual random acts of individuals. (A)*

Seeing the patient in the context of a system of care also provided a context for ICP, helping to reveal the interprofessional interdependence inherent in even seemingly simple patient care scenarios,

> *They don’t know how many people touch a patient and so now they’re starting to figure out the complexity of one patient’s care. (B)*

## Discussion

The study participants articulated an integrated approach to teaching both quality improvement and interprofessional collaborative practice where ICP is backgrounded as a critical pre-requisite and QI is foregrounded as a vector for engendering interprofessional competencies, with systems thinking as a mediating interface. The interaction and synergy between these two fields are crucial to maximizing their impact and the most powerful and sustained results generally follow from a combination of both QI and interprofessional interventions. ^44,45,68–72^ This is what we found at MU, a practice we exhort other institutions to follow. Additionally, reviewers have voiced their frustration at not being able to tease out the exact impact of ICP, for example, in part because most of the successful interventions do not sufficiently clarify curricular details and often include bundled strategies, frequently with a focus on what are arguably QI-related skills. ^73–76^ The MU case and these reviews bolster our argument that these fields are profoundly intertwined, further challenging their continuing separation.

Interestingly, MU faculty described interprofessional competencies from the perspective of attitudes and skills, while defining quality improvement competencies in terms of knowledge and skills. Existing competency domain statements articulate knowledge, skills, and attitudes for both fields, leaving us to puzzle at this finding. Perhaps it is evidence of interdependency between the two fields, especially when they are explicitly pursued together in program design. While QI has long held to Deming’s inclusion of psychology of human work and change as a key domain of improvement knowledge, relational skills required for managing the psychology of change in an intensely hierarchical and complex multi-professional environment like healthcare are perhaps unique and better described by ICP.^34^ Alternatively, when talking about their teaching, faculty may have assigned the attitude competencies to ICP and the knowledge competencies to QI as a matter of convenience.

While studies in health professional education, especially in medicine, have named and tried to understand the impact of the hidden curriculum often described as implicit and unintentional from the institution or program perspective, little attention has been paid to the advantages of a deliberate approach to an implicit curriculum, i.e. backgrounding.^77,78^ Faculty in this study describe an intentional approach to an implicit curriculum around interprofessional collaborative practice designed to attenuate the stance that these competencies can be taken for granted as already known or practiced. By backgrounding ICP, educators can potentially build competence in interprofessional collaborative practice while inviting learners to focus on a quality problem or challenge.

The background-foreground concept resonates with the notion of quality improvement and other complex problems as boundary objects, i.e. “physical, abstract, or mental object[s] that [serve] as a focal point in collaboration enabling parties to represent, transform and share knowledge.”^79(p1805)^ Boundary objects, for example clinical care pathways, can serve as flexible yet concrete means to bridge hierarchies, foster collective learning and growth, and promote a common culture between distinct professional identities. ^80–82^ Yet it is likely not sufficient to have a boundary object at play. We must also pay attention to the underlying interpersonal dynamics which mediate the teams’ interaction with a problem. Issues like psychological safety are critical and antecedent to preventing errors and improving performance.^83–85^ While acting as a boundary object in the foreground, a quality improvement initiative offers faculty and learners an opportunity to span professional silos. At the same time, interprofessional competencies operating in the background are crucial to the team’s success. The two work better synergistically.

Finally, systems thinking emerged as a mediating interface, relevant both to quality improvement in efforts to solve complex system problems and specific to interprofessional collaborative practice as it helps faculty and learners provide and enhance individual care for those with complex needs.^86–88^ The relationship between QI and ICP as described by faculty at MU brings the definition of interprofessional education to life – “learning with, from, and about each other” to solve real system and patient-specific problems with a view to improving care and outcomes for individuals and populations.^4^

## Methodological Considerations and Further Research

The results derived from the study of a crucial 10-year period at MU are a first step in developing an integrated ICP and QI competency framework. This instrumental case study with a faculty group at MU has allowed us to generate new knowledge on the how and why of QI-ICP education and contribute to theoretical generalization. ^89^ It is important to note that the respondents are only a subset of the MU faculty who are involved in this program of teaching and learning, and participation from faculty outside of medicine and nursing was limited. Additionally, while the study period was over a decade ago, we confirmed recently with a subset of the study participants that work to maintain and improve their approach to teaching QI-ICP content continues to this day. Further research would benefit from involving an expanded range of interprofessional faculty, including rehabilitation sciences, social work, support services, pharmacy, and health care administration. Including more of these other faculty may have allowed us to uncover other aspects of MU’s unique program that would illustrate other concepts and principles important to the integration of ICP and QI. Another institutional setting altogether might also yield new insights or provide deeper understandings. Finally, a concrete next step might be to use the suggested competency domains and sub-themes presented here to develop and test specific competency statements across several health professions education settings.

## Conclusion

The MU case demonstrates the potential for teaching synergies when faculty address quality improvement and interprofessional collaborative practice competencies with an integrated approach that leverages connections, while also respecting distinctions. For health professions education programs looking to improve the effectiveness and efficiency of their curricular approach to these fields, it may be fruitful to consider ICP as background and QI as foreground, remembering that without each other, ICP risks losing meaning and QI risks losing impact.

## Data Availability

All data produced in the present work are contained in the manuscript

## Acknowledgments

The authors wish to thank the faculty participants at the University of Missouri for giving generously of their time and wisdom.

## Ethical approval

Ethical approval for this study was waived by the MU IRB, April 27, 2012, IRB#1202415

## Footnotes

b In 2015, the Institute of Medicine became the National Academy of Medicine, but these reports were published prior to that transition.

c Multiprofessional teams include team members from different health and social care professions working in parallel rather than interactively.^39^

d An instrumental case study focuses on broad concepts, in this case the integration of QI and IPC as elucidated through the experience at MU from 2003-13. That is in contrast with an intrinsic case study, where the subject of interest is the case itself, i.e., the specifics of the experience at MU.

e While observing one session, we identified an additional participant and so one participant was involved in only a stimulated-recall interview, which was conducted post-observation.

